# Why didn’t people get vaccinated against COVID-19? Results from a nationwide survey among Mexican adults

**DOI:** 10.1101/2024.01.17.24301326

**Authors:** Dagmara Wrzecionkowska, Christopher R. Stephens, Juan Pablo Gutiérrez Reyes

**Affiliations:** Universidad Nacional Autónoma de México (UNAM)

**Keywords:** COVID-19, unvaccinated, reasons for non-vaccination

## Abstract

**Objective:** To explore the reasons for not getting vaccinated against COVID-19.

**Material and methods:** In October 2021, a nationwide structured telephone survey (disproportionate stratified sampling) was conducted regarding COVID-19 pandemics, including vaccination experience. To examine associations between inoculation and other characteristics, the chi-square test and logistic regression analysis were applied.

**Results:** Out of 3 126 adults, 68% reported complete vaccination and 21% only the first dose, while 11% remained unvaccinated. Non-vaccination was associated with being younger, male, without a partner, low socioeconomic level, and no previous diagnosis of hypertension, obesity or diabetes. Furthermore, the non-vaccinated were less likely to have tested for COVID-19, and more likely to consider COVID-19 as low severe and not real compared with the vaccinated. Using logistic regression models: place of residence, marital status, educational level, age, BMI, testing for COVID-19, and the perception of COVID-19 (severe and real) were significant predictors of non-vaccination. The predominant reasons for not getting vaccinated were: 63% “external barriers” (e.g., not being able to attend an appointment), and 37% “internal motives” (e.g., “vaccine does not work”).

**Conclusions:** The causes of non-vaccination against COVID-19 are related to both social and geographical determinants. Addressing external barriers is necessary in order to promote equity in vaccination. Reviewing the results in the context of earlier studies on the willingness to vaccinate, the gap between intention and vaccination is notable.

## Introduction

The COVID-19 pandemic has accounted for 6.7 million deaths worldwide up to January 2023. Mexico is one of the most affected countries, with the highest case-fatality ratio (deaths per 100 confirmed cases).^1^. In the case of epidemics, vaccination is considered one of the most effective ways to combat a disease at the population level.^2^ In Mexico, the SARS-CoV-2 vaccination program began in December 2020. In October 2021, 78% of adults had received at least one dose (the period of data collection for this study), leaving 22% as unvaccinated.^3^

Vaccine Hesitancy (VH) defined as “the reluctance or refusal to vaccinate despite the availability of vaccines”, was recognized by the WHO in 2019 as one of the top ten health threats.^2^ According to the WHO vaccine advisory group, VH is affected by: convenience, confidence and complacency. Convenience refers to the ease of obtaining the service, considering external barriers, such as vaccine availability, affordability and quality. Confidence relates to the degree of trust placed in the safety and effectiveness of the vaccine, competence, and reliability of those who apply it and the policy makers. Complacency includes the perception of risk level from contracting a disease vs. risks associated with getting vaccinated. Despite of including convenience, the Strategic Advisory Group of Experts Working Group (SAGE WG) has noted that the scope of vaccine hesitancy excludes external factors on the public health level associated with vaccine accessibility.^4^

In the case of the SARS-CoV-2 vaccine, its acceptance varies across countries. Among 81 studies reviewed by Shakeel at al.^5^ acceptance levels ranged from 97% in Ecuador to 21% in Lebanon. Lazarus at al.^6^, based on a survey conducted in June 2020, reported a world average of 71.5%, and, in Mexico, 75%. A better understanding of the reasons behind VH would allow the design of more effective vaccination campaigns. Although several studies have already estimated the levels and factors associated with VH worldwide, and specifically in Mexico, the majority of these were conducted prior to SARS-Cov-2 vaccine availability. The objective of this study was to identify the level of COVID-19 non-vaccination in Mexico and the predictors of remaining unvaccinated. The secondary objective was to identify reasons given for not getting vaccinated.

## Materials and methods

A cross-sectional study was conducted analyzing data collected between September and October 2021 via a telephone-survey of individuals living in Mexico. Telephone numbers were randomly selected using prefixes to approximate the geographical distribution of the population in the country (non-proportional stratified random sampling). Sample size (n) was estimated based on the formula 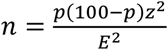, p representing the percentage occurrence of a condition, E the percentage maximum error required, and z the value corresponding to level of confidence required. For this study the following values were applied: p=50%, z=1.96, E=2%, assuming a response rate of 50%, so a total of 6000 numbers were selected to ensure 3000 complete interviews.

The questionnaire was designed *ad hoc* based on other instruments, with variables grouped into four sections:

1. **Socio-demographics and socio-economic status (SES)** age, sex, educational level, marital status, region of residence.
2. **Vaccination against COVID-19 status** first dose only, complete vaccination, non-vaccination; type of vaccine received, reasons behind not getting vaccinated with multiple response options.
3. **COVID-19** testing and report of the disease from the individual and other in her/his household. with items: Have you had any tests done to find out if you have COVID-19? Have you had a positive COVID-19 test result? Have you been diagnosed with COVID-19 by a doctor? Do you think you have had COVID-19? Has anyone in your household other than you had COVID-19? And perception of COVID-19 with items: Do you think COVID-19 is real? with response options: yes/no; How serious do you consider COVID-19 on a scale from 1 to 5? 5 meaning very severe.
4. **Other health status items:** comorbidities (previous diagnosis of diabetes and hypertension from a health professional); Body Mass Index (BMI), calculated with self-reported weight and height; smoking; hand washing frequency on the day of the interview and the number of facemasks owned.

Individuals were grouped into the following geographical regions, defined to assure similar numbers of participants per region: North, State of Mexico, Mexico City, Other central states, and South.^7^ A socioeconomic status (SES) index with 5 categories was calculated as the sum of five items: having a desktop computer, a laptop, a car, paid TV and internet connection at home.

Informed consent was obtained from each participant. The protocol for the research reported in this paper was approved by the Ethics Committee of the School of Medicine, National Autonomous University of Mexico (FMED/CEI/PMSS/153/2023).

## Analysis

Data from the participants who provided complete information (n=3126) were analyzed with a chi-square test to compare all characteristics for the vaccinated and unvaccinated. Two logistic models were built using forced-entry logistic regression. The first model assessed the associations between getting vaccinated with a first dose against COVID-19 (dependent variable, with response options yes/no) and other variables. In the second model the dependent variable was not getting vaccinated due to internal barriers vs. external barriers. The same independent variables were applied as in the first model. There were no issues of multicollinearity between the independent variables. Adjusted odds ratios (AORs) and 95% confidence intervals (CIs) were calculated adjusting for age (introduced as a continuous variable), marital status (nominal dichotomous), residence area (nominal dummy coded, with Mexico City as a reference), educational level (ordinal), sex (nominal dichotomous) and socio economic status (ordinal variable). In the first model, only the first four variables were significant, and in the second only age and education.

All statistical analyses were performed with IBM ® SPSS version 25, with a *p* value of 0.05 taken as the level of significance.

## Results

A total of 3,126 adults completed the survey (65% women), from 18 to 92 years old (mean 43 +/-16). Of those, 56% were married or lived together, 32% had education level below secondary or secondary, 34% high school, 34% undergraduate or graduate. The sample covered all 32 states of Mexico, with a predominant representation from Mexico City 24% and State of Mexico 19%.

A total of 1294 individuals (41% of the sample) reported having been tested for COVID-19, with 444 (34% out of those tested) obtaining a positive result. Reported prevalence of COVID-19 in the total sample was 30% (n=923), this included those with a positive test result, diagnosed by a doctor and/or those who considered having had COVID-19.

Regarding COVID-19 vaccination, 68% (n=2125) reported having received two doses (or a single-shot vaccine like Cansino) and 21% (n=654) only a first dose, while 11% (n=347) were unvaccinated. The two most frequently applied vaccines were AstraZeneca (42%) and Pfizer (28%), followed by Sinovac (11%), Sputnik V (9%), and Cansino (6%). With respect to the participants’ beliefs, only 3% considered COVID-19 as unreal and only 4% considered it little severe. The number of facemask that participants owned on average was 23 (+/-54), and the frequency of hands washing on the day of the interview on average was 10 (+/-8).

The sample consisted predominantly of those with overweightedness and obesity (65%), the group with normal weight represented 34%, and those with underweightedness 2%. 15% indicated having been diagnosed with diabetes and 18% with hypertension (Table I).

**Table I.**
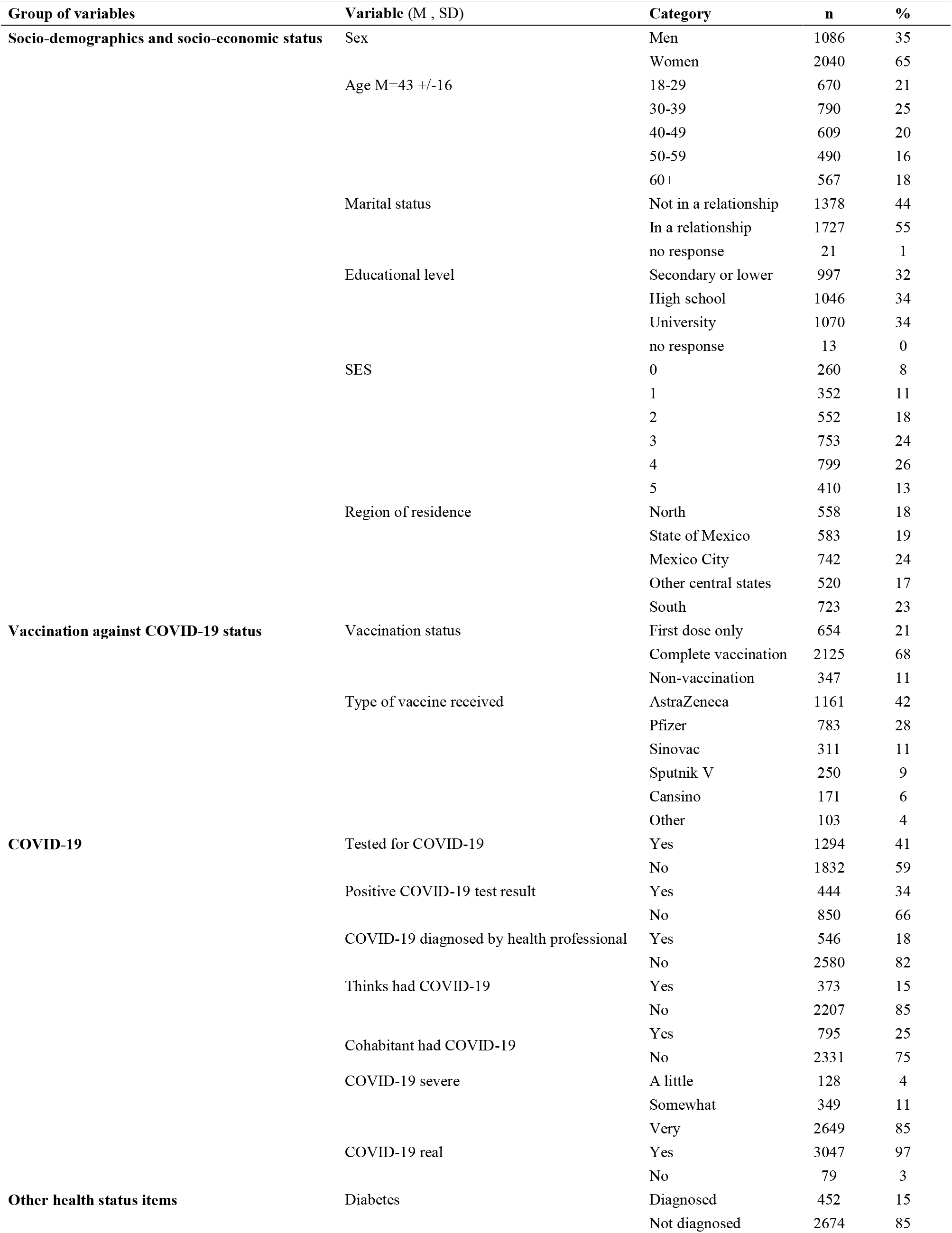
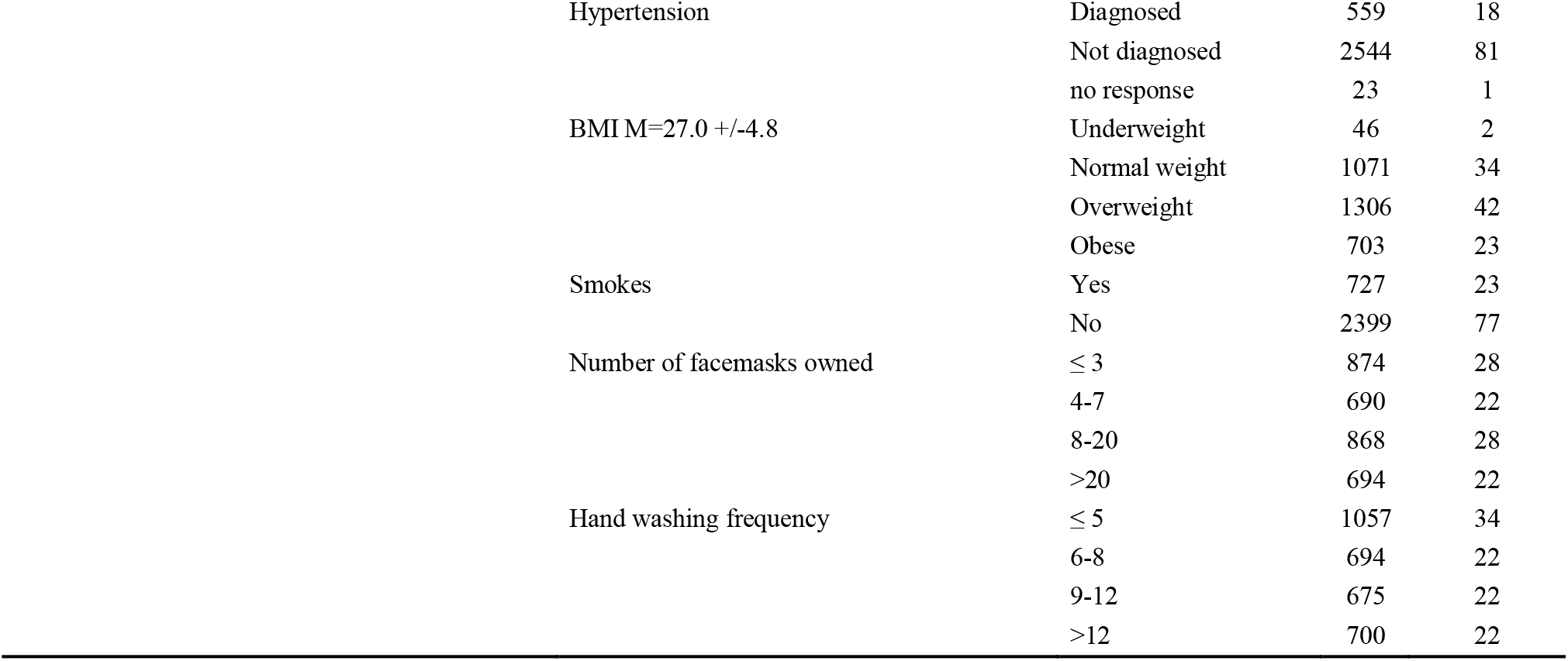
Descriptive characteristics for the whole sample n= 3126

| Group of variables | Variable (M , SD) | Category | n | % |
| --- | --- | --- | --- | --- |
| **Socio-demographics and socio-economic status** | Sex | Men | 1086 | 35 |
|  |  | Women | 2040 | 65 |
|  | Age M=43 +/-16 | 18-29 | 670 | 21 |
|  |  | 30-39 | 790 | 25 |
|  |  | 40-49 | 609 | 20 |
|  |  | 50-59 | 490 | 16 |
|  |  | 60+ | 567 | 18 |
|  | Marital status | Not in a relationship | 1378 | 44 |
|  |  | In a relationship | 1727 | 55 |
|  |  | no response | 21 | 1 |
|  | Educational level | Secondary or lower | 997 | 32 |
|  |  | High school | 1046 | 34 |
|  |  | University | 1070 | 34 |
|  |  | no response | 13 | 0 |
|  | SES | 0 | 260 | 8 |
|  |  | 1 | 352 | 11 |
|  |  | 2 | 552 | 18 |
|  |  | 3 | 753 | 24 |
|  |  | 4 | 799 | 26 |
|  |  | 5 | 410 | 13 |
|  | Region of residence | North | 558 | 18 |
|  |  | State of Mexico | 583 | 19 |
|  |  | Mexico City | 742 | 24 |
|  |  | Other central states | 520 | 17 |
|  |  | South | 723 | 23 |
| **Vaccination against COVID-19 status** | Vaccination status | First dose only | 654 | 21 |
|  |  | Complete vaccination | 2125 | 68 |
|  |  | Non-vaccination | 347 | 11 |
|  | Type of vaccine received | AstraZeneca | 1161 | 42 |
|  |  | Pfizer | 783 | 28 |
|  |  | Sinovac | 311 | 11 |
|  |  | Sputnik V | 250 | 9 |
|  |  | Cansino | 171 | 6 |
|  |  | Other | 103 | 4 |
| **COVID-19** | Tested for COVID-19 | Yes | 1294 | 41 |
|  |  | No | 1832 | 59 |
|  | Positive COVID-19 test result | Yes | 444 | 34 |
|  |  | No | 850 | 66 |
|  | COVID-19 diagnosed by health professional | Yes | 546 | 18 |
|  |  | No | 2580 | 82 |
|  | Thinks had COVID-19 | Yes | 373 | 15 |
|  |  | No | 2207 | 85 |
|  | Cohabitant had COVID-19 | Yes | 795 | 25 |
|  |  | No | 2331 | 75 |
|  | COVID-19 severe | A little | 128 | 4 |
|  |  | Somewhat | 349 | 11 |
|  |  | Very | 2649 | 85 |
|  | COVID-19 real | Yes | 3047 | 97 |
|  |  | No | 79 | 3 |
| **Other health status items** | Diabetes | Diagnosed | 452 | 15 |
|  |  | Not diagnosed | 2674 | 85 |

|  |  |  |  |
| --- | --- | --- | --- |
| Hypertension | Diagnosed | 559 | 18 |
|  | Not diagnosed | 2544 | 81 |
|  | no response | 23 | 1 |
| BMI M=27.0 +/-4.8 | Underweight | 46 | 2 |
|  | Normal weight | 1071 | 34 |
|  | Overweight | 1306 | 42 |
|  | Obese | 703 | 23 |
| Smokes | Yes | 727 | 23 |
|  | No | 2399 | 77 |
| Number of facemasks owned | ≤ 3 | 874 | 28 |
|  | 4-7 | 690 | 22 |
|  | 8-20 | 868 | 28 |
|  | >20 | 694 | 22 |
| Hand washing frequency | ≤ 5 | 1057 | 34 |
|  | 6-8 | 694 | 22 |
|  | 9-12 | 675 | 22 |
|  | >12 | 700 | 22 |

A comparison between vaccinated and unvaccinated individuals is reported in Table II. Being younger, male, without a partner, low SES, residing in the State of Mexico or Other Central State, not having comorbidities, smoking, not considering COVID-19 a real or severe disease, and adopting COVID-19 protective behaviors less frequently were all statistically significant predictors for not getting vaccinated.

**Table II.**
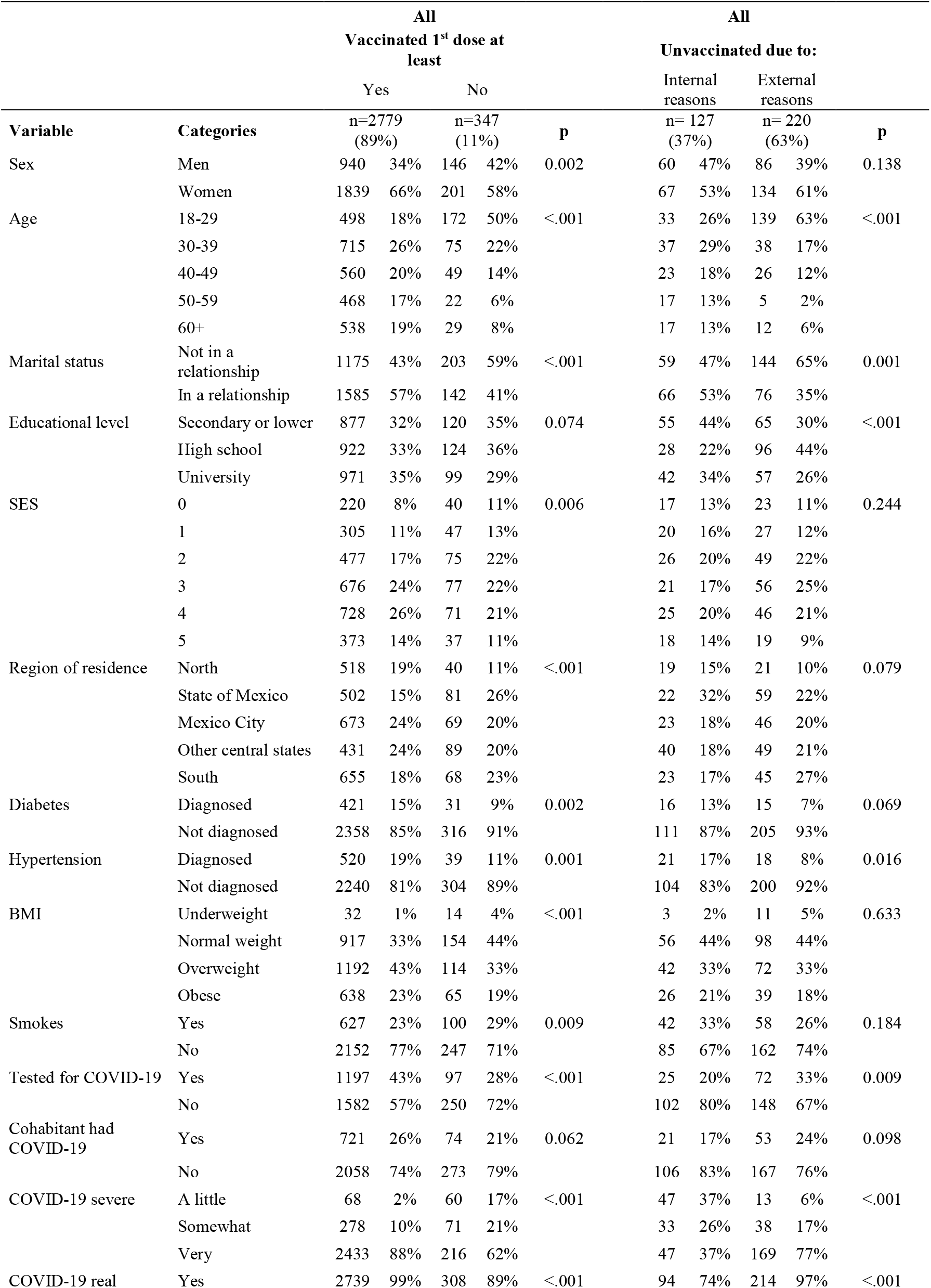
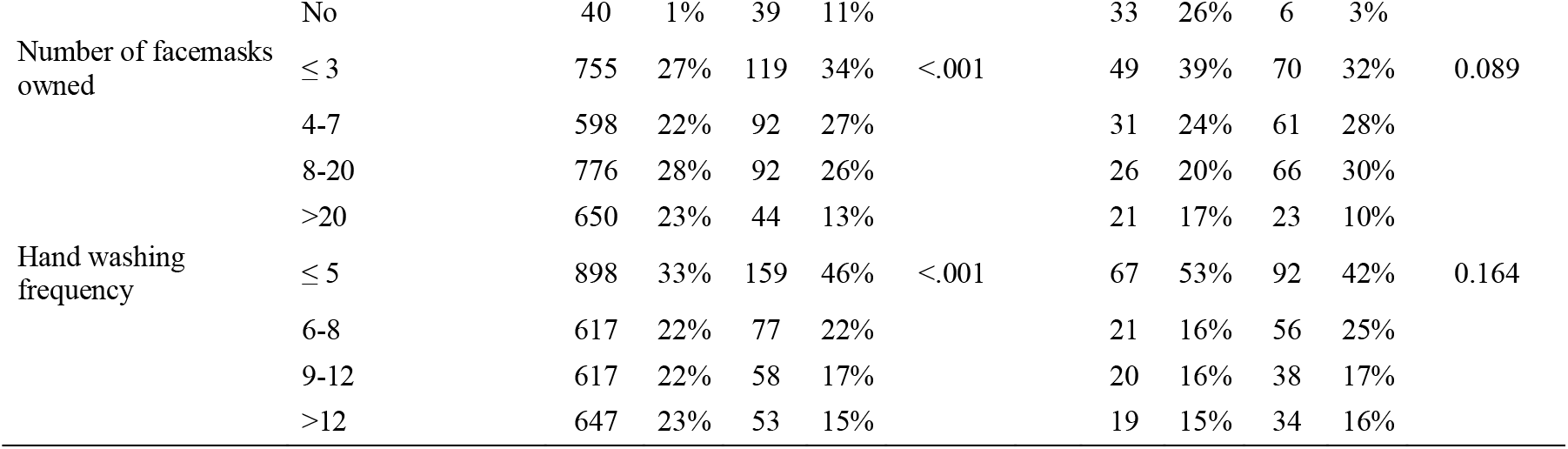
Comparison between: 1. vaccinated with one or more doses and the unvaccinated; and 2. **unvaccinated** due to **internal** barriers vs. **external** barriers, applying a chi squared test.

|  |  | All<br>Vaccinated 1<sup>st</sup> dose at<br>least |  |  |  |  | All<br>Unvaccinated due to: |  |  |  |  |
| --- | --- | --- | --- | --- | --- | --- | --- | --- | --- | --- | --- |
|  |  | Yes |  | No |  |  | Internal<br>reasons |  | External<br>reasons |  |  |
| Variable | Categories | n=2779<br>(89%) |  | n=347<br>(11%) |  | P | n= 127<br>(37%) |  | n= 220<br>(63%) |  | P |
| Sex | Men | 940 | 34% | 146 | 42% | 0.002 | 60 | 47% | 86 | 39% | 0.138 |
|  | Women | 1839 | 66% | 201 | 58% |  | 67 | 53% | 134 | 61% |  |
| Age | 18-29 | 498 | 18% | 172 | 50% | <.001 | 33 | 26% | 139 | 63% | <.001 |
|  | 30-39 | 715 | 26% | 75 | 22% |  | 37 | 29% | 38 | 17% |  |
|  | 40-49 | 560 | 20% | 49 | 14% |  | 23 | 18% | 26 | 12% |  |
|  | 50-59 | 468 | 17% | 22 | 6% |  | 17 | 13% | 5 | 2% |  |
|  | 60+ | 538 | 19% | 29 | 8% |  | 17 | 13% | 12 | 6% |  |
| Marital status | Not in a relationship | 1175 | 43% | 203 | 59% | <.001 | 59 | 47% | 144 | 65% | 0.001 |
|  | In a relationship | 1585 | 57% | 142 | 41% |  | 66 | 53% | 76 | 35% |  |
| Educational level | Secondary or lower | 877 | 32% | 120 | 35% | 0.074 | 55 | 44% | 65 | 30% | <.001 |
|  | High school | 922 | 33% | 124 | 36% |  | 28 | 22% | 96 | 44% |  |
|  | University | 971 | 35% | 99 | 29% |  | 42 | 34% | 57 | 26% |  |
| SES | 0 | 220 | 8% | 40 | 11% | 0.006 | 17 | 13% | 23 | 11% | 0.244 |
|  | 1 | 305 | 11% | 47 | 13% |  | 20 | 16% | 27 | 12% |  |
|  | 2 | 477 | 17% | 75 | 22% |  | 26 | 20% | 49 | 22% |  |
|  | 3 | 676 | 24% | 77 | 22% |  | 21 | 17% | 56 | 25% |  |
|  | 4 | 728 | 26% | 71 | 21% |  | 25 | 20% | 46 | 21% |  |
|  | 5 | 373 | 14% | 37 | 11% |  | 18 | 14% | 19 | 9% |  |
| Region of residence | North | 518 | 19% | 40 | 11% | <.001 | 19 | 15% | 21 | 10% | 0.079 |
|  | State of Mexico | 502 | 15% | 81 | 26% |  | 22 | 32% | 59 | 22% |  |
|  | Mexico City | 673 | 24% | 69 | 20% |  | 23 | 18% | 46 | 20% |  |
|  | Other central states | 431 | 24% | 89 | 20% |  | 40 | 18% | 49 | 21% |  |
|  | South | 655 | 18% | 68 | 23% |  | 23 | 17% | 45 | 27% |  |
| Diabetes | Diagnosed | 421 | 15% | 31 | 9% | 0.002 | 16 | 13% | 15 | 7% | 0.069 |
|  | Not diagnosed | 2358 | 85% | 316 | 91% |  | 111 | 87% | 205 | 93% |  |
| Hypertension | Diagnosed | 520 | 19% | 39 | 11% | 0.001 | 21 | 17% | 18 | 8% | 0.016 |
|  | Not diagnosed | 2240 | 81% | 304 | 89% |  | 104 | 83% | 200 | 92% |  |
| BMI | Underweight | 32 | 1% | 14 | 4% | <.001 | 3 | 2% | 11 | 5% | 0.633 |
|  | Normal weight | 917 | 33% | 154 | 44% |  | 56 | 44% | 98 | 44% |  |
|  | Overweight | 1192 | 43% | 114 | 33% |  | 42 | 33% | 72 | 33% |  |
|  | Obese | 638 | 23% | 65 | 19% |  | 26 | 21% | 39 | 18% |  |
| Smokes | Yes | 627 | 23% | 100 | 29% | 0.009 | 42 | 33% | 58 | 26% | 0.184 |
|  | No | 2152 | 77% | 247 | 71% |  | 85 | 67% | 162 | 74% |  |
| Tested for COVID-19 | Yes | 1197 | 43% | 97 | 28% | <.001 | 25 | 20% | 72 | 33% | 0.009 |
|  | No | 1582 | 57% | 250 | 72% |  | 102 | 80% | 148 | 67% |  |
| Cohabitant had COVID-19 | Yes | 721 | 26% | 74 | 21% | 0.062 | 21 | 17% | 53 | 24% | 0.098 |
|  | No | 2058 | 74% | 273 | 79% |  | 106 | 83% | 167 | 76% |  |
| COVID-19 severe | A little | 68 | 2% | 60 | 17% | <.001 | 47 | 37% | 13 | 6% | <.001 |
|  | Somewhat | 278 | 10% | 71 | 21% |  | 33 | 26% | 38 | 17% |  |
|  | Very | 2433 | 88% | 216 | 62% |  | 47 | 37% | 169 | 77% |  |
| COVID-19 real | Yes | 2739 | 99% | 308 | 89% | <.001 | 94 | 74% | 214 | 97% | <.001 |
|  | No | 40 | 1% | 39 | 11% |  | 33 | 26% | 6 | 3% |  |
| Number of facemasks owned | ≤ 3 | 755 | 27% | 119 | 34% | <.001 | 49 | 39% | 70 | 32% | 0.089 |
|  | 4-7 | 598 | 22% | 92 | 27% |  | 31 | 24% | 61 | 28% |  |
|  | 8-20 | 776 | 28% | 92 | 26% |  | 26 | 20% | 66 | 30% |  |
|  | >20 | 650 | 23% | 44 | 13% |  | 21 | 17% | 23 | 10% |  |
| Hand washing frequency | ≤ 5 | 898 | 33% | 159 | 46% | <.001 | 67 | 53% | 92 | 42% | 0.164 |
|  | 6-8 | 617 | 22% | 77 | 22% |  | 21 | 16% | 56 | 25% |  |
|  | 9-12 | 617 | 22% | 58 | 17% |  | 20 | 16% | 38 | 17% |  |
|  | >12 | 647 | 23% | 53 | 15% |  | 19 | 15% | 34 | 16% |  |

**Table III.** Model characteristics

|  | **Model 1** |  |  |  | **Model 2** |  |  |  |
| --- | --- | --- | --- | --- | --- | --- | --- | --- |
|  | DV got vaccinated (yes/no) |  |  |  | DV did not get vaccinated due to internal vs external reasons |  |  |  |
| Prediction for the class: | Unvaccinated |  |  |  | Unvaccinated due to internal reasons |  |  |  |
| Model reliability | $\chi^2=370.530$, df=9, p<0.001 | | | | $\chi^2=126.516$, df=4, p<0.001 | | | |
| Hosmer and Lemeshow test | $\chi^2=4.046$, df=8, p=.853 | | | | $\chi^2=4.933$, df=8, p=.765 | | | |
| Variance explained |  |  |  |  |  |  |  |  |
| Cox & Snell | 0.11 |  |  |  | 0.31 |  |  |  |
| Nagelkerke | 0.23 |  |  |  | 0.42 |  |  |  |
| Total correct predictions | 0.89 |  |  |  | 0.79 |  |  |  |
| Prediction class 1 | 98.9% of vaccinated |  |  |  | 89.9% of unvaccinated due to external barriers |  |  |  |
| Prediction class 2 | 11.7% of unvaccinated |  |  |  | 60.8% of unvaccinated due to internal barriers |  |  |  |
|  | B | Standard Error | Wald | df | p value | OR | 95 CI |  |
|  |  |  |  |  |  |  | Inferior | Superior |
| **Model 1** |  |  |  |  |  |  |  |  |
| Region of residence |  |  |  |  |  |  |  |  |
| Other Central States | 0.67 | 0.16 | 18.27 | 1 | 0.00 | 1.95 | 1.44 | 2.65 |
| State of Mexico | 0.40 | 0.16 | 6.64 | 1 | 0.01 | 1.49 | 1.10 | 2.03 |
| Age | -0.05 | 0.01 | 88.62 | 1 | 0.00 | 0.95 | 0.94 | 0.96 |
| Educational level | -0.27 | 0.07 | 16.76 | 1 | 0.00 | 0.76 | 0.67 | 0.87 |
| Marital Status *Single* | 0.37 | 0.13 | 8.00 | 1 | 0.01 | 1.45 | 1.12 | 1.88 |
| BMI | -0.03 | 0.01 | 4.68 | 1 | 0.03 | 0.97 | 0.94 | 1.00 |
| No test for COVID-19 | 0.54 | 0.14 | 15.09 | 1 | 0.00 | 1.71 | 1.30 | 2.24 |
| COVID-19 is unreal | 1.07 | 0.30 | 12.95 | 1 | 0.00 | 2.92 | 1.63 | 5.23 |
| COVID-19 is severe | -0.50 | 0.06 | 60.56 | 1 | 0.00 | 0.61 | 0.54 | 0.69 |
| **Model 2** |  |  |  |  |  |  |  |  |
| Age | 0.07 | 0.01 | 38.24 | 1 | 0.00 | 1.07 | 1.05 | 1.09 |
| Educational level | 0.48 | 0.15 | 10.57 | 1 | 0.00 | 1.61 | 1.21 | 2.15 |
| COVID-19 is unreal | 1.24 | 0.55 | 5.07 | 1 | 0.02 | 3.46 | 1.17 | 10.22 |
| COVID-19 is severe | -0.83 | 0.14 | 36.24 | 1 | 0.00 | 0.44 | 0.33 | 0.57 |

Note: DV dependent variable

Within the group of the unvaccinated (n=347), 63% (n= 220) of the individuals reported “external” barriers for non-vaccination. Those barriers included not being able to attend a vaccination appointment, while 37% (n=127) reported “internal” reasons, such as beliefs that the vaccine was harmful or ineffective, or there was a lack of interest (Figure I).

**Figure I.**
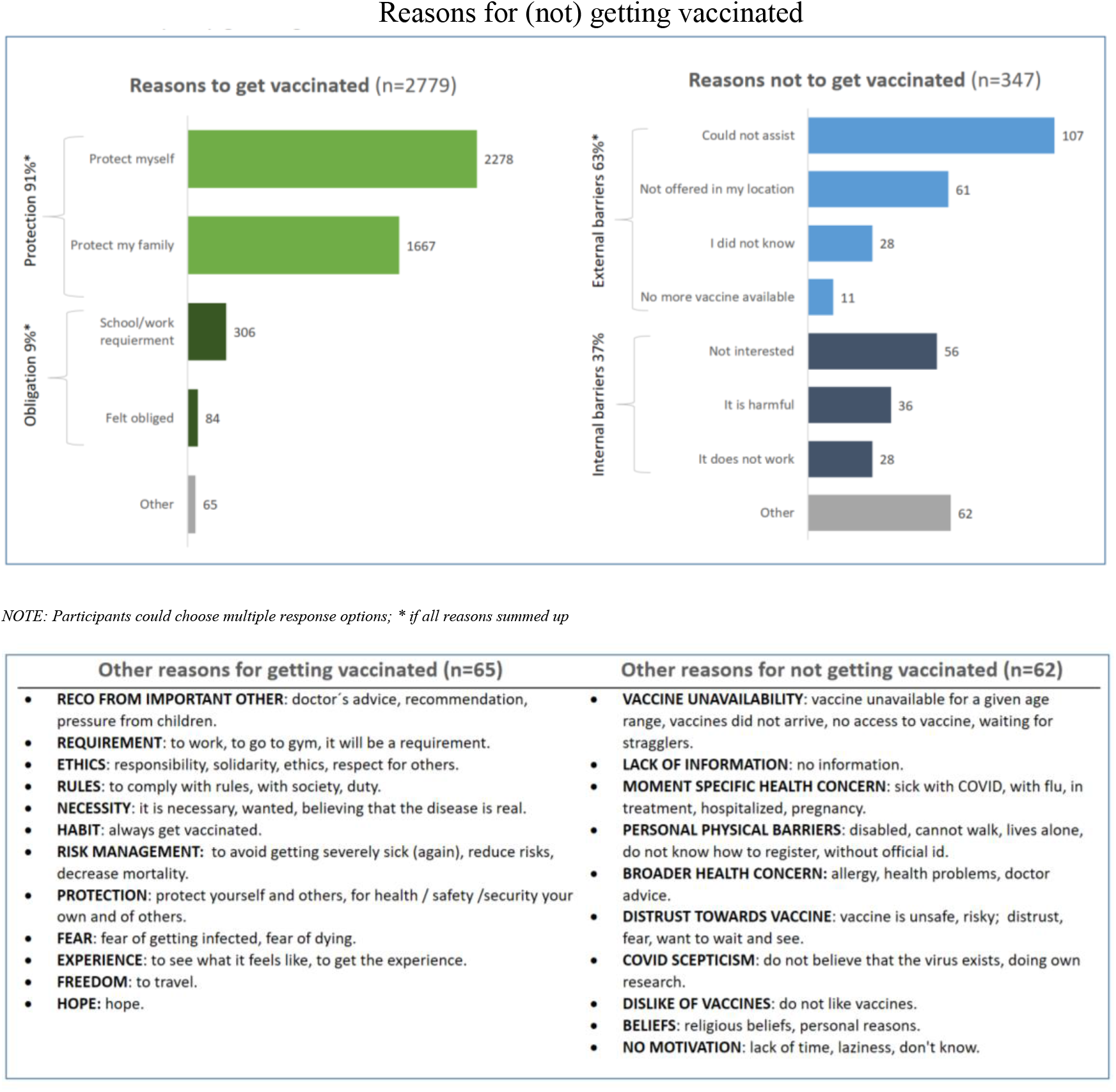
Reasons for (not) getting vaccinated

Not getting vaccinated for internal reasons vs. external ones was associated with being between 50-59 years of age; with educational level no higher than secondary or having a university degree; residing in northern states; not living with someone who had COVID-19, and perceiving COVID-19 as not severe or not real (Table II).

When asked for reasons for not getting vaccinated, participants could elaborate if they considered that none of the provided response options applied to them. Within the additional responses several could fit the provided options. Additional reasons for not getting vaccinated encompassed e.g., health problems such as allergies, being pregnant, skepticism as to the virus existence, personal/religious beliefs. Some provided specific reasons for not being able to assist, such as not having a valid ID, not knowing how to register, or not being able to walk - see Figure I.

In logistic regression analysis Model 1 with getting vaccinated as a dependent variable (yes/no response options) and Model 2 with not get vaccinated due to internal vs. external reasons as a dependent variable, were significantly reliable and adequately fit the data. See Table II for the model characteristics and AORs. Region of residence (Other central state OR 1.95, 95 CI 1.44, 2.65; State of Mexico OR 1.49, 95 CI 1.10, 2.03) and COVID-19 related beliefs (COVID-19 unreal OR 2.92, 95 CI 1.63, 5.23; COVID-19 severe OR 0.61, 95 CI 0.54, 0.69) were the strongest significant predictors, followed by marital status (OR 1.45, 95 CI 1.12, 1.87), getting tested for COVID-19 (OR 1.71, 95 CI 1.30, 2.23), and education level (OR 0.76, 95 CI 0.67, 0.87). Residing in the State of Mexico or Other Central States were associated with not getting vaccinated. Those who considered COVID-19 as not real were almost 3 times more likely not to get vaccinated; and for every unit increase in perceiving COVID-19 as severe there was a 39% reduction in the probability to not get vaccinated. Singles were more likely by 45% not to get vaccinated, and with every unit increase in educational level there was a 24% reduction in the probability not to get vaccinated. Younger age (OR 0.95, 95 CI 0.94, 0.96) and lower BMI (OR 0.97, 95 CI 0.94, 1.00) were also associated with not getting vaccinated. In Model 2 only age (OR 1.07, 95 CI 1.05, 1.09), education level (OR 1.61, 95 CI 1.21, 2.15) and factors related to the perception of COVID-19 were statistically significant (COVID-19 unreal OR 3.46, 95 CI 1.17, 10.22; COVID-19 severe OR 0.43, 95 CI 0.33, 0.57).

## Discussion

In this study, 11% of respondents indicated not having been vaccinated, with the reasons for not getting vaccinated divided into two groups: external barriers that accounted for 63% of the reasons mentioned and internal barriers 37%. **External barriers** encompassed vaccine limited availability and factors, such as not being able to attend an appointment or not having information about vaccination, which could indicate that either the information was not provided, or that the person did not pay attention. **Internal barriers** included considering the vaccine as harmful or ineffective and not being interested.

Although the percentage of unvaccinated individuals is lower than the percentage reported by the government (22%) in the comparable period, this difference could be attributed to the methodology of telephone survey. Also, our data included a larger share of women, who were more likely to get vaccinated, and inhabitants of Mexico City (24%), the region with a higher level of vaccination compared to other regions of Mexico.

When combining the vaccinated with the unvaccinated due to external barriers (i.e. individuals that may be vaccinated if having access) the level of vaccination would reach 97%, in line with the prior research conducted in Mexico where 3% answered that they would not get vaccinated.^8^

Significant predictors for not getting vaccinated were, in the order of their strength: region of residence, COVID-19 related beliefs, marital status, previous testing for COVID-19, education level, age and BMI.

Regarding the **area of residence**, the highest prevalence of vaccination was registered in northern states (93%) and the lowest in the State of Mexico (86%) and Other central states (83%), results that are consistent with those from the ENSANUT (national survey conducted by Secretaria de Salud, called Encuesta Nacional de Salud y Nutrición^7^). Lower vaccination in the Other central states could be related to access to the vaccine. As reported for Colombia, Ecuador, and Venezuela structural barriers to vaccination, specifically the difficulties of getting vaccinated in rural areas, may decrease vaccination rates.^9^ Beyond accessibility, availability factors e.g., lack of vaccination infrastructure, services, cold chain supply and distance-related factors, were mentioned in a review conducted in Latin America prior to the COVID pandemic as barriers to getting vaccinated.^10^

In this study 45% reported being **single**, and this group was almost 1.5 times less probable to be vaccinated compared to couples, although being single is associated with younger age, so age could be an underlying confounder for a lower vaccination rate in this group.

Lower **education level** was associated with higher rates of non-vaccination. Yet, analyzing the group of unvaccinated due to internal reasons, the percentages were similar at or below high school level and at the university level, pointing to a nonlinear relationship between the variables. In Mexico some studies, conducted prior to the vaccine availability, or at its early stage, indicated no effect of education level on the willingness to get vaccinated^11^ (data was collected between December 2020 and February 2021). Others suggested that those without a high school diploma or with a vocational-technical degree show a significantly high proportion of low VH and a low proportion of no VH^8^ (data was collected between June and July, 2021,). The relation between education and VH requires further research.

The **age** group 18-29 had the highest percentage of the unvaccinated compared to the older groups, a result that is contrary to some studies realized prior to or at early stages of the vaccine availability, where the youngest group expressed the highest levels of vaccine acceptance and the oldest showed highest hesitancy.^8,11-13^ Also, earlier reviews conducted in Latin America, including Mexico, that assessed VH for other vaccines indicated that advanced age was associated with vaccination concerns and refusal.^10^ However, vaccination prevalence for SARS-CoV-2 shows quite a contrary picture. Evidence from Mexico indicates that there are more unvaccinated among young adults, than within the oldest age group.^7^ Higher vaccination rates in older age groups can be explained via earlier eligibility for getting vaccinated, their greater risk of severe consequences, including higher mortality, which was mediatized, and past experience with vaccines.

Higher **BMI** was associated with higher rates of vaccination. Similar results were found in the UK.^14^ In Mexico, Ramonfaur et al.^12^ found that important variables associated with a higher vaccine acceptance included having any comorbidity (a web-based nationwide survey was conducted in early December 2020). Quite early in the development of the pandemic, obesity was identified as one of the key factors that increased the risk of being hospitalized or dying from COVID-19.^15^

Not taking the **COVID-19 test** was other significant predictor for not getting vaccinated. This association seems to have been less studied, yet we could assume that getting tested and vaccinated fits a profile of a person who is more concerned about their health and might perceive themselves at risk of COVID-19. On the other hand, those who do not **perceive COVID as severe** and do not perceive themselves in danger of getting sick might consider the prevention measures less necessary. After controlling for all the other variables, perceiving COVID-19 as being unreal (3% of the sample) and little or not at all severe (4% of the sample) were strong predictors of not getting vaccinated. Karlsson et al.^16^ found that those who perceived COVID-19 as a severe disease were also slightly more willing to receive a COVID-19 vaccination.

Some **limitations** of the study need to be mentioned. Sampling was random yet non-proportionally stratified. Collecting data via telephone might have skewed the profile of participants. Some variables, that were indicated in other studies as significant predictors for non-vaccination, were not included: vaccination history, health insurance, trust in government and the healthcare system. Participants were presented with a list of potential reasons for not being vaccinated, and although we allowed for other reasons to be mentioned, there may be still other motives that were not identified.

## Conclusions

Decision making regarding whether or not to get vaccinated is a complex phenomenon, with multiple factors involved. The main ones identified in this study included area of residence, socio-demographics (marital status, education level, age), health related reasons (comorbidities) and beliefs related to risk perception of COVID-19. Although this study was cross-sectional, reviewing the obtained results in the context of earlier studies, provided some evidence regarding the dynamic character of VH. Also, comparing results of this study, that focused on vaccine uptake, with studies that investigated acceptance or intention to get vaccinated prior to vaccine availability, highlight the gap between intention and behavior, calling for further research. Furthermore, this study adds to the evidence that accessibility barriers and lack of information play an important role in Mexico, that represents the global south. This study provides further evidence that structural solutions are necessary in Latin America, specifically in Mexico, to ensure –and maintain—high levels of vaccination.

## Data Availability

All data produced in the present study are available upon reasonable request to the authors.

